# Stimulation-related increases in power spectral density covary with clinical evidence of overstimulation during deep brain stimulation for pediatric dystonia

**DOI:** 10.1101/2025.03.05.25322884

**Authors:** Madelyn Pascual, Pritha Bisarad, James Kelbert, Sarah Chinander, Rose Gelineau-Morel, Carolina Gorodetsky, Angela L. Hewitt, Travis Larsh, Nicole Lucente, Jennifer O’Malley, Terrence D. Sanger, Lauren van der Werf, Jason S. Hauptman, Francisco A. Ponce, Michael C Kruer, John A Thompson

**Affiliations:** Pediatric Movement Disorders Program, Department of Neurology, Barrow Neurological Institute, Phoenix Children’s, Phoenix, Arizona, USA; Departments of Cellular & Molecular Medicine, Child Health, Neurology and Translational Neuroscience and Program in Genetics, University of Arizona College of Medicine – Phoenix, Phoenix, Arizona, USA; Department of Rehabilitation Therapy, Phoenix Children’s, Phoenix, Arizona, USA; Department of Neurology, Children’s Mercy Medical Center, Kansas City, MO, USA; Hospital for Sick Children, Toronto, Ontario, Canada; Department of Neurology, University of Rochester, Rochester, NY, USA; Division of Neurology, Cincinnati Children’s Hospital Medical Center, Cincinnati, Ohio, USA and Department of Pediatrics, University of Cincinnati College of Medicine, Cincinnati, Ohio, USA; Department of Pediatrics, Stanford University, Palo Alto, CA, USA; Department of Neurology, Children’s Hospital of Orange County, Los Angeles, CA, USA; Department of Neurosurgery, Barrow Neurological Institute, Phoenix Children’s, Phoenix, Arizona, USA; Department of Neurosurgery, Barrow Neurological Institute, Dignity Health, Phoenix, Arizona, USA; Program in Biomedical Informatics, College of Health Solutions and Programs in Neuroscience and Molecular & Cellular Biology, School of Life Sciences, Arizona State University, Tempe, Arizona, USA; Departments of Neurology & Neurosurgery, Anschutz Medical Campus, University of Colorado – Aurora, Aurora, Colorado, USA

## Abstract

**BACKGROUND:** Dystonia patients undergoing deep brain stimulation (DBS) often require individualized stimulation settings. While effective settings reduce dystonia, excessive stimulation can worsen symptoms. Assessing DBS effects during office visits is challenging, as clinical changes can be delayed hours to days.

**OBJECTIVES:** We evaluated whether local field potentials (LFPs) could serve as an acute biomarker of excessive stimulation in dystonia patients.

**METHODS:** Real-time LFP band power and dystonia severity were quantified and compared during sequential changes in stimulation amplitude.

**RESULTS:** Dystonia worsening was temporally associated with clinically evident and statistically significant increases in LFP band power during in-office DBS programming sessions.

**CONCLUSIONS:** Although increased LFP band power correlated with clear clinical worsening in these patients, we anticipate that not all patients with dystonia will have such immediate signs of worsening. Increased LFP band power during incremented stimulation amplitude may represent a biomarker for patients at-risk of manifesting delayed clinical worsening.

## INTRODUCTION

Dystonia is the third most prevalent movement disorder behind Parkinson Disease and essential tremor^1^. Dystonia is characterized by sustained (tonic) or intermittent (phasic)^2^ muscle contractions causing abnormal, often repetitive, movements or postures^3^ and/or non-velocity dependent hypertonia^4^. Dystonia, particularly its phasic elements, can be quite variable, with sensory and/or emotional triggers^5^ and fluctuating severity, manifesting in its most extreme form as status dystonicus. Dystonia is highly associated with chronic pain and impaired function^6^.

Available dystonia treatments include an array of medications, injection therapies, and surgeries. Management of dystonia is complicated by the variable responses that are often observed from patient-to-patient, even among those with similar clinical presentations or the same genetic condition^7^. Despite the many medical treatment options for dystonia, available options focus on symptom suppression and medication alone is often unable to adequately control dystonia^8^. Both botulinum toxin and phenol injections can ameliorate the symptoms of dystonia, and botulinum toxin is often recommended as first-line treatment for patients with focal or segmental dystonias^9^. However, for patients with severe and/or generalized dystonia, even high-dose botulinum injections are often unable to adequately control their symptoms^9^. Surgical treatments, including ventral rhizotomy, lesional approaches, intrathecal baclofen pump placement, and deep brain stimulation (DBS) can be valuable options for patients with severe, refractory, and/or generalized dystonia^10^. For appropriate patients, DBS can lead to dramatic reductions in symptom severity, exemplified by patients with DYT1, who on average experience a ∼70-80% reduction in symptoms^11,12^. For pediatric patients, early utilization of DBS may be associated with improved outcomes, potentially preventing musculoskeletal deformity^13^.

Nevertheless, although some patients experience life-changing benefits from DBS, others respond only modestly. This variability in DBS outcomes from patient-to-patient represents a major challenge for clinicians^14^. Anatomically, a ‘sweet spot’ for electrode placement for dystonia patients has been identified in the globus pallidus^15^ and optimal lead placement could help reduce variability in outcomes. Some of the variability in DBS outcomes is also related to etiology, but even among patients with the same mutation in the same gene different responses can be encountered^16^, indicating that additional factors beyond lead placement and etiology influence outcomes. One potential contributor to variable outcomes for dystonia patients using DBS is the latency between stimulation changes and clinical effects. In comparison to patients with Parkinson Disease or tremor who often demonstrate symptom reductions in the office setting, clinical responses to changes in DBS stimulation are often delayed days to weeks, sometimes months, for patients with dystonia^17,18^.

Another factor likely contributing to the variable response seen is the difficulty inherent in identifying both under- and over-stimulation. Dystonia patients typically require multiple programming sessions with a graded response to stimulation^19^. Historically, DBS settings have thus been changed during office visits, guided by both patient report and repeated neurological examinations, supported by video recordings and standardized rating scale scores used to monitor a patient’s response to stimulation. However, given that dystonia is characteristically variable throughout the course of the day, fluctuating with changes in emotional state, physical health, sensory stimulation, and both passive and active movement^20^, it can be challenging to rely solely on in-clinic evaluations, where patients with dystonia may experience heightened levels of stress. Finally, the traditional unidirectional (clinician to patient) approach to programming based on observation and report can be particularly challenging given that many pediatric dystonia patients may have a limited ability to communicate verbally or even with the support of augmentative communication technology such as eye gaze systems. We describe our recent observations in pediatric dystonia patients with DBS devices, reporting on a relationship between stimulation settings, local field potential (LFP) characteristics, and dystonia severity, providing evidence that states of ‘overstimulation’^21,22^ have a neurophysiologic correlate with potential clinical utility.

## PATIENTS & METHODS

We recorded real time continuous local field potentials (Active Streaming mode) data with video from 3 patients with generalized dystonia during DBS programming sessions. The patients and/or parents provided written informed assent/consent as applicable (Phoenix Children’s IRB #15-080) for publication, including representative videos. Data from each patient’s Medtronic Percept device were extracted as JSON files and analyzed using MATLAB v.2024b (Mathworks, Natick, Massachusetts, USA) (**see Supplemental Material**).

## RESULTS

For each patient, stepwise increases in DBS current amplitude were implemented during programming sessions. As current was serially increased, each patient demonstrated objective evidence of ‘overstimulation’ (worsening of clinical dystonia during a continuously monitored Active Streaming session). In each case, patients demonstrated a threshold effect with onset of clinical overstimulation, followed by further worsening of clinical dystonia with successive increases in amplitude (see **Supplemental Material & Videos**). Continuing to increase stimulation amplitude led to a visually recognizable increase in LFP power spectral density (**Figure 1A-H**) that correlated with time-locked and video-based measures of dystonia severity (Burke-Fahn-Marsden (BFM) dystonia rating scale).

**Figure 1:**
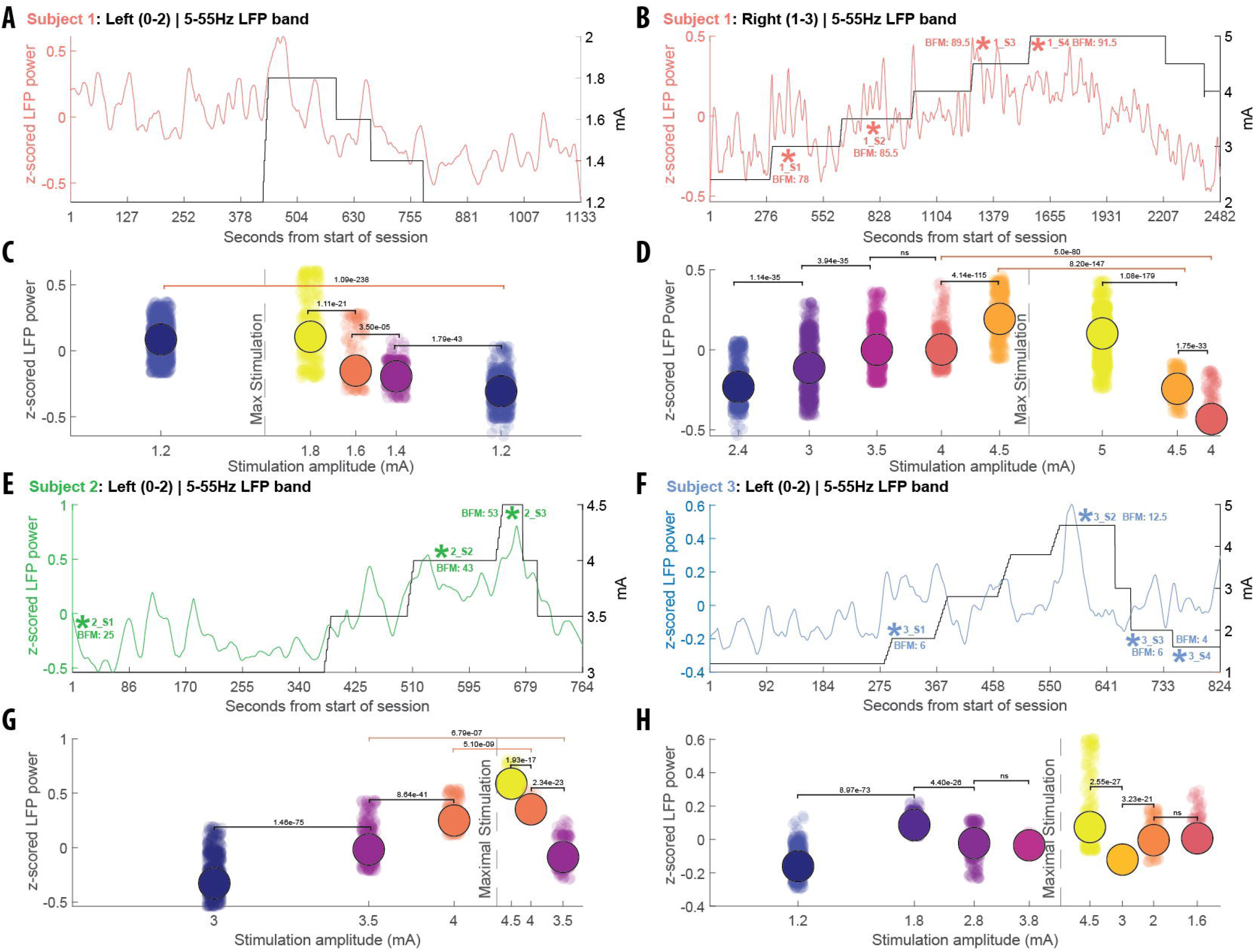
Individual subject (red = Subject 1; green = Subject 2; blue = Subject 3) LFP band power responses to changes in stimulation magnitude. Supplemental videos and Burke-Fahn-Marsden (BFM) scores are indicated. Power spectral density (5-55 Hz) for left (A) and right (B) GPi during current ramp-based stimulation changes. Left (C) and right (D) LFP power changes per stimulation amplitude bin for Subject 1 (black connecting lines denote comparisons within either pre- or post-maximum stimulation amplitude and red connecting lines denote comparisons between mirrored amplitudes across pre- and post-maximum amplitude). (E) and (G) show LFP band power response to stimulation for Subject 2. (F) and (H) show LFP response to stimulation for Subject 3. Wilcoxon rank sum test was used to evaluate significant differences in LFP power across successive stimulation amplitudes and between the pre- and post-maximum stimulation amplitude conditions. Both amplitudes and stimulation conditions were evaluated with post-hoc comparisons.

For each subject, we calculated BFM scores from available video and compared power spectral density (PSD) using Wilcoxon rank sum tests before vs after successive steps in the current ramp (**Figure 1**). We found that once a certain undefined stimulation threshold was exceeded, PSD increased in a highly significant manner. This threshold varied from one patient to the next. Furthermore, we found that once PSD had significantly increased from one step to the next, this pattern continued, correlating with successive worsening of dystonia, until maximal stimulation was reached. In cases where current was then serially decreased, this was accompanied by a successive improvement in dystonia. Interestingly, when LFP band power was later compared for the same current before and after maximal stimulation, PSD was significantly lower for post-maximal current steps.

We then performed a linear mixed-effects analysis to assess the relationship between current, PSD, and dystonia severity as captured by BFM score. We found a significant main effect of current amplitude, indicating that BFM ratings were higher with greater stimulation if PSD was also increased (**Figure 2**). Of potential clinical relevance, PSD magnitude was largely driven by user-selected beta band power (**Supplemental Figure 1**), suggesting that clinical monitoring allows for visual discrimination and selection of the most informative frequency bands to track and correlate with patient response. LFP band power did not significantly predict an increase in BFM on its own (LFP = -4.881), but the combined effects of stimulation amplitude and LFP band power interactions were significant (mA:LFP = 8.725), indicating that the effect of stimulation on BFM depended on mean LFP power. Substantial inter-individual differences were observed (subject intercept SD = 37.34), indicating variability across participants in baseline BFM that corroborated clinical impressions.

**Figure 2:**
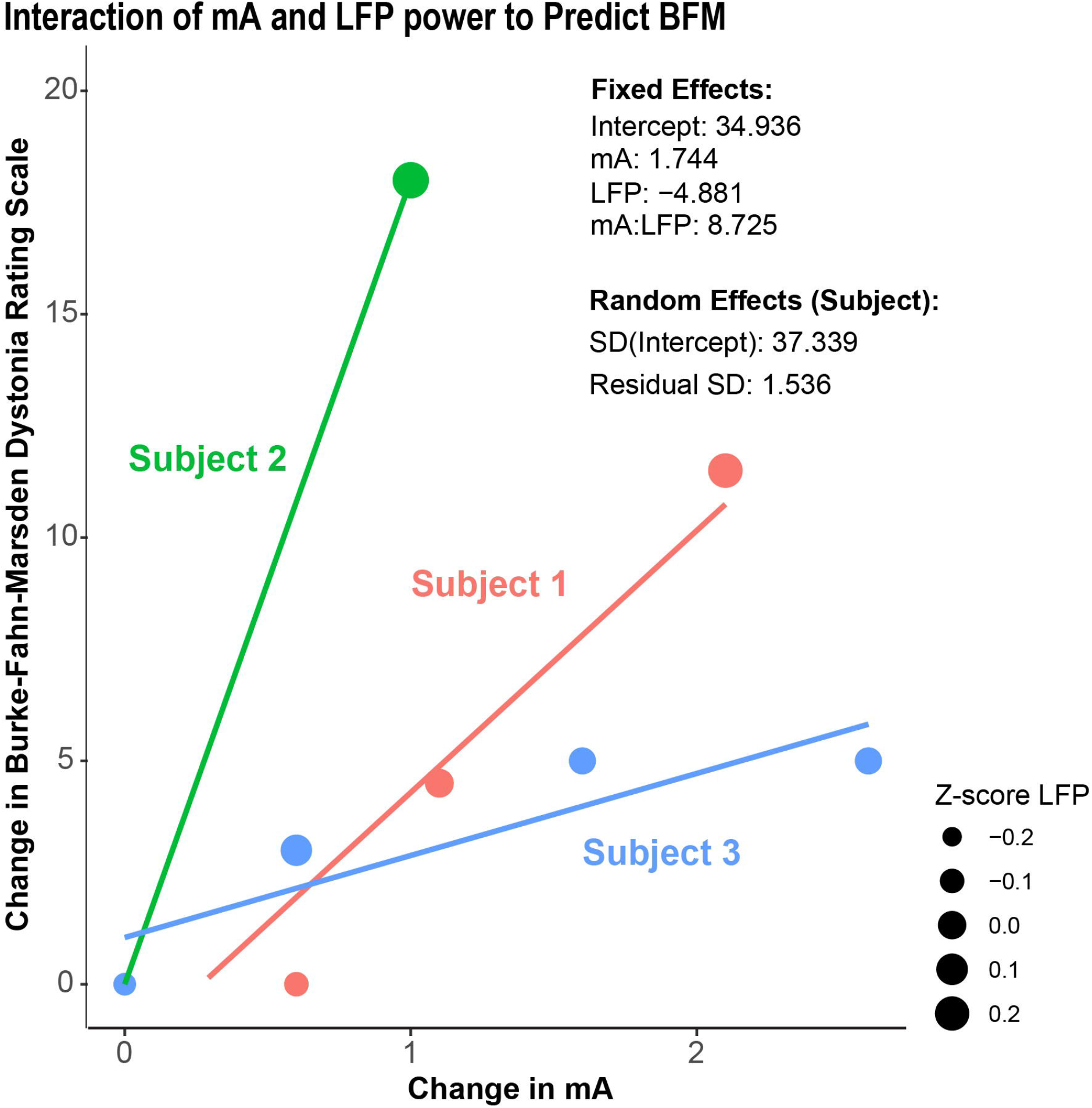
Linear mixed model analysis assessing the combined ability of stimulation current and mean LFP power to predict changes in dystonia severity (BFM scores) in all 3 subjects. All subjects values indicate an increase in BFM scores covaries with greater interaction between stimulation magnitude and mean LFP power. Fixed effects statistics (baseline intercept of 34.936) estimate a positive relation between stimulation and BFM (mA = 1.744) but a negative relation between mean power and BFM (LFP = -4.881). Overall statistics indicate a positive interaction between stimulation current and mean LFP power (mA:LFP = 8.725) suggesting that increased stimulation positively impacts mean LFP power leading to overall increase in BFM score. Random effects statistics (standard deviation of baseline intercept = 37.339) suggest significant variability in BFM baseline across the 3 subjects.

## DISCUSSION

Fundamentally, when DBS is beneficial, effective stimulation settings lead to a reduction in dystonia severity. Prior publications have shown that clinically beneficial DBS stimulation is also associated with a reduction in LFP PSD in the GPi. In our patients, mixed modeling showed that the converse is also true - when increased stimulation co-occurred with clinically evident increases in PSD magnitude, this was associated with a consistent worsening in clinical dystonia symptoms as captured by Burke Marsden Dystonia rating scale scores. Although overstimulation has been described clinically in patients^21,22^ knowledge of its neurophysiologic basis has been limited.

We found that reducing stimulation after a patient was clinically deemed to be exhibiting signs of overstimulation led to a highly significant reduction in PSD magnitude, also supporting a causal relationship between stimulation intensity and longitudinal PSD. Once a state of overstimulation was recognized, reducing stimulation intensity led to a concurrent decrease in dystonia severity, again supporting a direct relationship between stimulation intensity, PSD and dystonia severity. Taken together, these findings suggest that stimulation-related changes in PSD may be bidirectionally informative, with decreases in PSD correlating with *reduction* in clinical dystonia severity and *increases* in PSD correlating with *increases* in dystonia severity.

Of course, the utility of DBS in dystonia is based upon a therapeutic goal of using direct current stimulation to reduce dystonia. Our data indicates that more is not always better and suggests that dystonia patients may have a U-shaped response curve. In addition, our data indicates that each patient’s “optimal stimulation” state may differ. It is particularly interesting that Subjects 2 and 3 have the same mutation in the same gene causing their dystonia (DYT1) but that their stimulation requirements were nevertheless different.

Several important caveats also exist. Although overall we observed an increase in dystonia severity in association with an increase in PSD, these clinical observations required sufficient time for longitudinal evaluation. It is intriguing that each patient showed a lower PSD at a given stimulation current after a state of maximal stimulation had first been reached. This may represent a wash-in effect. Although this will require further study, if this phenomenon reproducibly occurs, there may be clinical value in titrating stimulation to the point of observable overstimulation, then serially decreasing stimulation to a more tolerable level - in some cases, this may allow stim to be pushed to a level that initially was not tolerated

Anecdotally, in other patients we have observed PSD increases without an appreciable increase in dystonia symptom burden. The clinical significance of these findings is unknown, but these observations are intriguing given that we know that for dystonia patients, benefits can be delayed days-weeks after changes to stimulation settings. Is the same true for overstimulation? Will such patients experience a delayed onset of clinical worsening (“catching up” to their LFP profiles)? Will their LFPs prove to be independent of their clinical symptoms? Or will they “acclimate” to these programming changes, experiencing a gradual decline in their PSD over time? It may be instructive to collect Chronic Sensing data for these patients to help answer these questions.

Our observations – and the follow up questions they prompt – highlight the challenges inherent in relying on continuous data to program a device such as a DBS. Findings to date indicate that moment to moment observations should be confirmed to represent part of a more sustained net effect. These observations also indicate that PSD may be an informative neurophysiologic biomarker of dystonia severity in the clinical setting, although our results will need to be further replicated and extended. Nevertheless, our findings indicate that continuously collected LFP data can serve as a valuable data stream to help guide clinical decision-making in real time.

## Data Availability

All data produced in the present study are available upon reasonable request to the authors

## Acknowledgements

We thank the patients and families for their support of this work. We acknowledge the support of Jumana Hoque in organizing patient data. We are grateful for the support of Phoenix Children’s Pediatric DBS coordinator Nicole Del Col.

## Financial disclosures for the previous 12 months

This work was supported by Phoenix Children’s Foundation Innovation Circle grant 102424-01 to MCK. ALH’s work is supported by funding from the Child Neurology Career Development Program K12 and Child Neurology Foundation (CNF) Pediatric Epilepsy Research Foundation (PERF) Shields Grant award. JAT’s work is supported by NIH UH3 NS113769. MCK’s work is supported by NIH R01 NS106298 and R01 NS127108. PB is supported by a Clinical Research Training Fellowship from the American Academy of Neurology. The authors declare there are no additional disclosures to report.

## Ethical Compliance Statement

The study was approved by the Phoenix Children’s IRB (#15-080). The patients and/or legal authorized representatives provided express written assent/consent for the analysis of their data and publication of findings, including videos. We confirm that we have read the Journal’s position on issues involved in ethical publication and affirm that this work is consistent with those guidelines.

## FIGURES

**Supplemental Figure:**
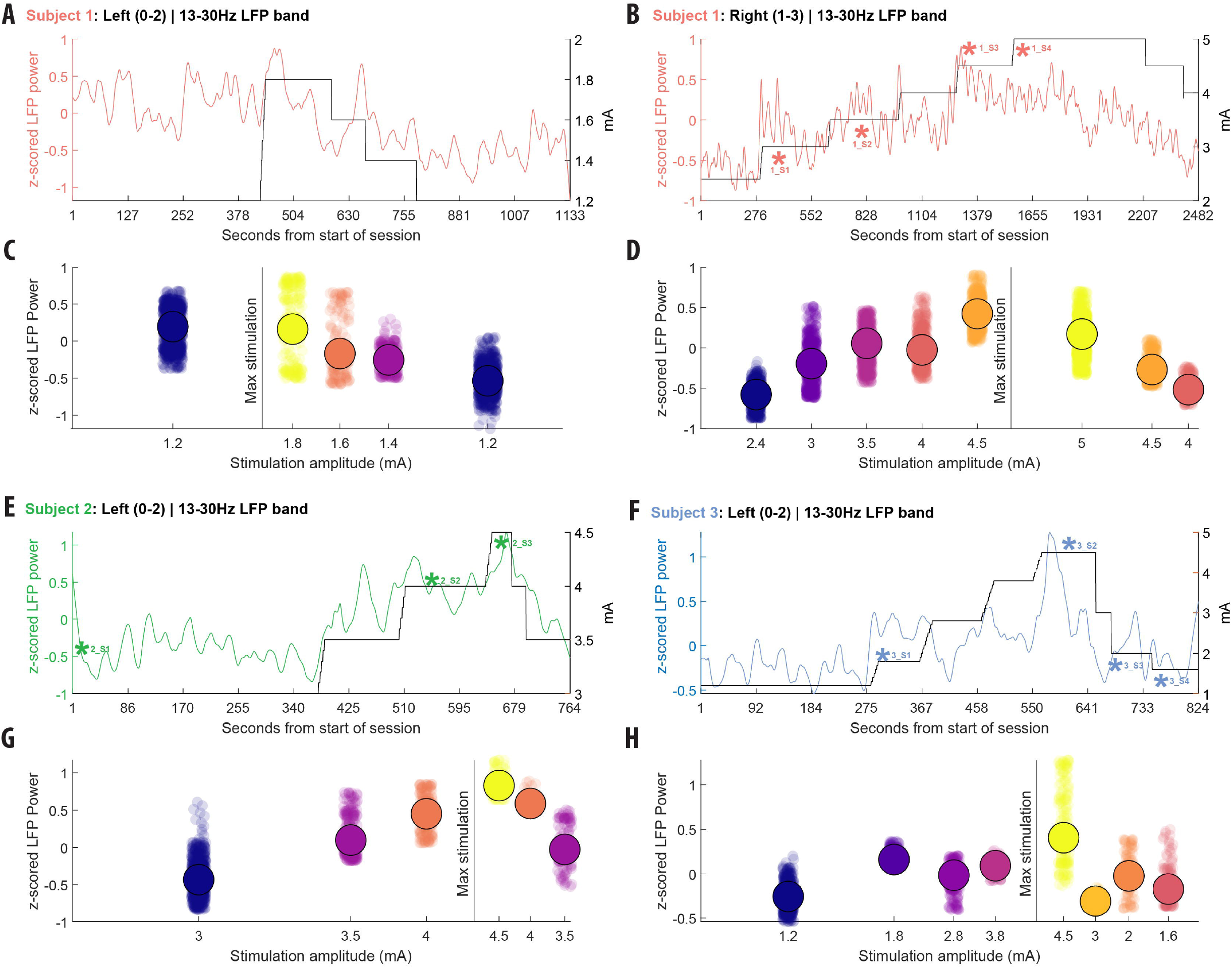
Individual subject (red = Subject 1; green = Subject 2; blue = Subject 3) LFP band power responses to changes in stimulation magnitude. Supplemental videos and Burke-Fahn-Marsden (BFM) scores are indicated. Beta band (13-30 Hz) power spectral density for left (A) and right (B) GPi during current ramp-based stimulation changes. Left (C) and right (D) LFP power changes per stimulation amplitude bin for Subject 1 (black connecting lines denote comparisons within either pre- or post-maximum stimulation amplitude and red connecting lines denote comparisons between mirrored amplitudes across pre- and post-maximum amplitude). (E) and (G) show LFP band power response to stimulation for Subject 2. (F) and (H) show LFP response to stimulation for Subject 3.

