## Supplemental Material for "Stimulation-related increases in power spectral density covary with clinical evidence of overstimulation during deep brain stimulation for pediatric dystonia"

**Supplemental Patients & Methods**

Bilateral globus pallidus pars interna-targeted Medtronic (Dublin, Ireland) leads were placed, coupled to an implantable pulse generator (IPG) with both LFP recording and streaming capability (Percept, Medtronic). Surgeries were without complications, and the patients tolerated the procedures well, returning to clinic for DBS activation 2-4 weeks after combined lead and IPG placement.

The Active Streaming function was utilized during office visits for the purpose of this study. Active Streaming captures LFPs with stimulation on. Specific LFP frequency bands (±2.5 Hz) of interest are visually represented during office visits on the device tablet, but data for the entire frequency spectrum was available for offline analysis. Videos were recorded during each programming session to document the patient’s symptoms and changes with programming.

Raw timeseries data were extracted from the JSON file (sampled at 250 Hz) and aligned with stimulation amplitude settings. For the LFP band power analysis, data were processed to remove EKG artifacts using the perceive toolbox (https://github.com/neuromodulation/perceive) and band-pass filtered from 5-55 Hz. Power was averaged in 1 second sample bins and normalized using the z-score method.

**Narrative description of each case**

Subject 1 was an 11-15 year-old female at the time of recording with generalized dystonic cerebral palsy GMFCS level V, microcephaly, and intellectual disability of unknown etiology. Her brain MRI disclosed only diminished cerebral volume. Her dystonia was recognized in infancy, starting in the left upper extremity then generalizing. She underwent bilateral GPi DBS during school age due to refractory generalized dystonia associated with chronic pain.

DBS leads were inserted into the globus pallidus interna (GPi) using the Leksell G-frame. Direct targeting was performed using proton density MR images to visualize the GPi, with the target being near the midcommisural plane where the laminations of the pallidum are most visible, inside the border between the external and internal segments of the globus pallidus, about 3-3.5 mm from the internal capsule. The deepest lead was placed beyond the target to the level of the optic tract. Stereotactic coordinates for the left GPi were targeted to (-15, 4, 0) with contact 2 measured at (-14.1, 2.9, -0.5), 0.8 mm from the targeted coordinates. For the right GPi (15, 3.5, -2) was targeted and contact 2 was found to be at (16.3, 5.7, 0.7), approximately 0.6 mm from the targeted coordinates.

The patient had cessation of her pain and improvement of her hand use but she continued to experience functionally disabling dystonia, so her implantable pulse generator was upgraded to a Percept PC system while a preteen. Her only medication was oxcarbazepine.

After several minutes of baseline data collection in Active Streaming mode, amplitude was first increased from 2.6 mA to 3.0 mA in the right GPi (1_S1). When stimulation amplitude was increased to 3.5 mA, higher power spectral density was evident and accompanied by clinical worsening of dystonia (1_S2). Stepwise increases to 4.0 mA and then to 4.5 mA were associated with more clearly evident worsening of the patient’s dystonia (1_S3), with increased repetitive movements and posturing of the left arm (0:00:38). At 5 mA, she experienced left hemifacial dystonia and persistent left arm dystonia with more frequent involuntary activation (1_S4) accompanied by a clinically appreciable increase in LFP peak magnitude. Stimulation amplitude was serially decreased, down to 1.8 mA (start of video) then to 1.6 mA (0:00:20) (1_S5).

Subject 2 was a 21-25 year-old young man with DYT1-associated heterozygous c.907_909del (p.Glu303del) deletion in *TOR1A* (paternally-inherited *TOR1A* heterozygous p.303del_E) generalized dystonia at the time of evaluation. He had prominent craniocervical involvement with whispering dysphonia. After normal pregnancy, delivery, and early development, he started to have gait abnormalities and lower limb dystonia during his elementary school years. His dystonia subsequently generalized. Cranial MRI showed an incidental finding of left mesial temporal cavernous malformation. This patient initially underwent DBS placement a few years after onset. He experienced good initial benefit but his dystonia control worsened in his teenage years, with painful daily dystonia. For this reason, his leads were reimplanted and his system was upgraded to a Medtronic Percept RC.

DBS leads were inserted into the globus pallidus interna (GPi) using the Leksell G-frame as described above. The deepest lead was again placed beyond the target to the level of the optic tract. Stereotactic coordinates targeted for the left GPi were (-21.5, 4.5, 0). Following lead placement, a stereotactic CT was obtained, showing the leads ~1.5 mm from the targeted coordinates (-19.9, 5, 1.3). For the right GPi, (21.25, 4.5, 0) was targeted, and stereotactic CT following lead placement showed the lead 0.8 mm from the targeted coordinates (20.7, 4, 0).

At the time of the recording, programming was still being actively adjusted. He was still able to walk independently but continued to experience painful neck and limb dystonia, hypophonia and dysarthria. During the programming session, a stepwise current ramp was implemented after several minutes of baseline data collection using Active Streaming mode. At 4.0 mA, the patient reported feeling involuntary activation of the right hemibody (2_S1). At 5.0 mA, he experienced worsening dystonia appreciable to the examiner, again on the right side of his body (2_S2). His LFPs showed higher magnitude peaks during clinical monitoring at this time.

Subject 3 was an 11-15 year-old female with DYT1 (heterozygous *de* *novo* *TOR1A* p.303del_E)-associated generalized dystonia with normal cranial MRI presenting as left foot dystonia during her elementary school years that subsequently generalized. Her dystonia was persistently worse on the left side of her body. She also experienced chronic daily pain related to her dystonia, struggled to walk and used a wheelchair for distances. She underwent placement of a Medtronic Percept RC DBS in bilateral GPi with significant improvement.

DBS leads were inserted into the globus pallidus interna (GPi) using the Leksell G-frame as described. The deepest lead was again placed beyond the target to the level of the optic tract. Stereotactic coordinates for the left GPi were (-18. 4.75, 0) while coordinates for the right GPi were (17.75, 4.5, -2). Following lead placement, a stereotactic CT was obtained, showing the leads 1.5 mm and 1.2 mm anterior to the left and right targets, respectively.

Baseline data was collected for several minutes, then a stepwise current ramp was implemented. At 1.6 mA, the patient reported worsening involuntary muscle activation of the right arm and hand and concurrently her LFP band peak magnitude appreciably increased (3_S1). When amplitude was increased to 2.8 mA (0:00:10) she had difficulty sustaining hand opening on the right and reported that her hand was closing involuntarily (3_S2). At 3.8 mA, she continued to experience tightness of the right hand and was unable to keep her hand open (3_S3). At 4.5 mA, she was still unable to sustain right hand opening (3_S4a) with a clinically-evident increase in LFP magnitude. She was also unable to open her hand fully and experienced cervical dystonia (3_S4b). Given these findings, the amplitude was serially decreased. At 2.0 mA, the patient reported her right hand felt better and she was able to open it more readily (3_S5). Amplitude was decreased further to 1.4 mA, and her hand control improved further (3_S6).
